# Circulating anti-Müllerian hormone levels in pre-menopausal women: novel genetic insights from a GWAS meta-analysis

**DOI:** 10.1101/2023.09.07.23295182

**Authors:** Natàlia Pujol-Gualdo, Minna K. Karjalainen, Urmo Võsa, Riikka K. Arffman, Reedik Mägi, Justiina Ronkainen, Triin Laisk, Terhi T. Piltonen

## Abstract

**Study question:** Can a genome-wide association study (GWAS) meta-analysis, including a large sample of young premenopausal women from a founder population from Northern Finland, identify novel genetic variants for circulating anti-Müllerian hormone (AMH) levels and provide insights into biological pathways and tissues involved in AMH regulation?

**Summary answer:** We identified six loci associated with AMH levels at *P* < 5 x 10^-8^, including the previously reported *MCM8*, *AMH* and *TEX41* loci, and three novel signals in or near *CHEK2*, *BMP4* and *EIF4EBP1*. Gene set enrichment analysis highlighted significant enrichment in renal system vasculature morphogenesis and tissue enrichment analysis ranks the pituitary gland as a top associated tissue.

**What is known already:** AMH is expressed by preantral and small antral stage ovarian follicles in women, and variation in age-specific circulating AMH levels has been associated with several health conditions. However, the biological mechanisms underlying the association between health conditions and AMH levels are not yet fully understood. Previous GWAS have identified loci associated with AMH levels in pre-menopausal women, but they were limited by small sample sizes or focused mostly on older pre-menopausal women.

**Study design, size, duration:** We performed a GWAS meta-analysis for AMH level measurements in 9,668 pre-menopausal women.

**Participants/materials, setting, methods:** We performed a GWAS meta-analysis in which we combined 2,619 AMH measurements (at age 31 years old) from a prospective founder population cohort (Northern Finland Birth Cohort 1966, NFBC1966) with a previous GWAS meta-analysis that included 7,049 pre-menopausal women (spanning age range 15-48). NFBC1966 AMH measurements were quantified using an automated assay (Elecsys® AMH Plus (Roche)). We annotated the genetic variants, combined different data layers to prioritise potential candidate genes, described significant pathways and tissues enriched by the GWAS signals, identified plausible regulatory roles using colocalization analysis and leveraged publicly available summary statistics to assess genetic and phenotypic correlations with multiple traits.

**Main results and the role of chance:** Three novel genome-wide significant loci were identified. One of these is in complete linkage disequilibrium with c.1100delC in *CHEK2*, which is found to be 4-fold enriched in the Finnish population compared to other European populations. We propose a plausible regulatory effect of some of the GWAS variants linked to AMH, as they colocalise with GWAS signals associated with gene expression levels of *BMP4*, *TEX41* and *EIFBP41*. Gene set analysis highlighted significant enrichment in renal system vasculature morphogenesis and tissue enrichment analysis ranked the pituitary gland as the top association.

**Large scale data:** The GWAS meta-analysis summary statistics will be available for download from the GWAS Catalog. Accession numbers will be provided upon publication.

**Limitations, reasons for caution:** This study only included women of European ancestry and the unavailability of sufficiently sized relevant tissue data in gene expression datasets hinders the assessment of potential regulatory effects in reproductive tissues.

**Wider implications of the findings:** Our results highlight the increased power of founder populations and larger sample sizes to boost the discovery of novel trait-associated variants underlying variation in AMH levels, which aided to characterise novel biological pathways and plausible genetic regulatory effects linked with AMH levels variation for the first time.

**Study funding / competing interest(s):** This work has received funding from the European Union’s Horizon 2020 research and innovation programme under the MATER Marie Sklodowska-Curie grant agreement No. 813707 and Oulu university scholarship foundation (N.P.-G.), Academy of Finland, Sigrid Jusélius Foundation, Novo Nordisk, University of Oulu, Roche Diagnostics (T.T.P). This work was supported by the Estonian Research Council grant 1911 (R.M.). J.R. was supported by the European Union’s Horizon 2020 research and innovation program under grant agreements No. 874739 (LongITools), 824989 (EUCAN- Connect), 848158 (EarlyCause) and 733206 (LifeCycle). U.V. was supported by the Estonian Research Council grant PRG (PRG1291). The NFBC1966 received financial support from University of Oulu Grant no. 24000692, Oulu University Hospital Grant no. 24301140, ERDF European Regional Development Fund Grant no. 539/2010 A31592.

## INTRODUCTION

Anti-Müllerian hormone (AMH) is a member of the transforming growth factor β (TGF-β) superfamily, which includes the bone morphogenic proteins (BMPs), growth differentiation factors, activins and inhibins. Despite owing its name to its classical role in male sexual differentiation, AMH is also expressed by the ovarian granulosa cells during the primary to small antral stage of follicle development (Weenen *et al*., 2004). In adult women, serum AMH levels decrease with age, with undetectable levels following menopause, signalling depletion of ovarian reserve (Finkelstei *et al*., 2020). As a result, AMH is primarily known as a serum marker for ovarian reserve. Previous studies have indicated that variations in age-specific circulating AMH levels are linked with several health conditions, including breast cancer (Ge *et al*., 2018) and polycystic ovary syndrome (PCOS) (Homburg and Crawford, 2014). Therefore, identifying genetic determinants of inter-individual variation in AMH measurements may offer valuable insight into their biological mechanistic effects and impact in health and disease beyond reproductive ageing.

Three previous genome-wide association studies have identified a few loci associated with AMH levels in pre-menopausal women (Schuh-Huerta *et al*., 2012; Ruth *et al*., 2019; Verdiesen *et al*., 2022). However, these studies were limited by small sample size (Schuh-Huerta *et al*., 2012) or focusing only or mostly on older pre-menopausal women (Ruth *et al*., 2019; Verdiesen *et al*., 2022). Since AMH decreases with age also the AMH variability decreases in older ages, and therefore there is less power to detect associations in older pre-menopausal women compared to younger pre-menopausal women. To address these limitations, our current work analyses data from a single time point measured in relatively young pre-menopausal women and combines this dataset with a previous meta-analysis (Verdiesen *et al*., 2022), to identify and characterise additional loci associated with AMH levels in pre-menopausal women.

The studies conducted so far have focused on European ancestry, mainly powered to discover associations with common genetic variants. Nonetheless, founder populations such as those found in northern Finland, represent a powerful resource to accelerate discovery of new biological mechanisms driven by low-frequency alleles that have risen to higher frequency due to unique demographic history (Kurki *et al*., 2023).

In the present study, we added 2,619 AMH measurements (at age 31 years old) from a prospective founder population cohort (Northern Finland Birth Cohort 1966, NFBC1966) to a previous GWAS meta-analysis of 7,049 pre-menopausal women (spanning age range 15- 48) (Verdiesen *et al*., 2022), reaching the largest sample size for assessing AMH variation in women from reproductive age to date (N=9,668). Beyond detecting three out of the four previously identified signals near *TEX41*, *MCM8* and *AMH*, we identified three novel signals near *EIF4EBP1*, *BMP4*, and *CHEK2*. In the *CHEK2* locus, the c.1100delC variant is enriched in the Finnish population and in complete linkage disequilibrium (r^2^=1) with the lead variant identified in that locus. Additionally, we tested regulatory effects of GWAS variants on specific genes and gene transcript expression in multiple tissues using colocalization analysis, evaluated gene set and tissue set enrichments, and estimated genetic correlations and phenotypic associations across multiple phenotypes, by leveraging publicly available summary statistics.

## METHODS

### Study design and setting

This study is based on the prospective population-based NFBC1966 study (University of Oulu, 1966, http://urn.fi/urn:nbn:fi:att:bc1e5408-980e-4a62-b899-43bec3755243). During 1966, 12231 children (of them 5,889 females) were born in the two northernmost provinces of Finland (covering 48% of Finnish territory). Originally, the study was set to evaluate early life factors on long-term health and work ability. The cohort population has been followed up at four different time points: 1, 14, 31 and 46 years of age (set by the cohort centre). Comprehensive questionnaires on female health and clinical examinations with biological data collection were performed at ages 31 and 46 years.

The detailed cohort description and follow up protocol has been previously published (Nordström *et al*., 2022). Briefly, in 1997 (the 31-year follow-up) women living in the Northern Finland area or in the Helsinki metropolitan area (n=4074 women) were invited to a clinical examination, in which 3,127 (77%) women participated. The present study includes 2,619 women with AMH and genetic data available at 31 years old (median=31.1, interquartile range (IQR)=30.9-31.4).

For the meta-analysis, publicly available summary statistics from an independent second study evaluating AMH variations across 7,049 pre-menopausal women were used (Verdiesen *et al*., 2022). Publicly available summary statistics were downloaded from the GWAS catalog (https://www.ebi.ac.uk/gwas/publications/35274129) under accession number GCST90104596. This study included data from the AMH GWAS meta-analysis by Ruth *et al*. (n=3,334) (Ruth *et al*., 2019), which meta-analysed four studies, and three cohorts which were additionally analysed and joined to the latter, conforming the most recent meta-analysis (n=7,049) (Verdiesen *et al*., 2022). The cohort details from the current meta-analysis (n=9,668) can be found in Supplementary Table 1. More details of cohort description of the studies included can be found in (Verdiesen *et al*., 2022). All studies had received ethical approval from institutional ethics committees.

### AMH measurement in NFBC1966

Serum samples were drawn in 1997 (the 31-year follow up), sealed and stored at −20℃ since then. Serum AMH concentrations were measured using the automated Elecsys® electrochemiluminescence immunoassay on a Cobas E411 analyser according to the manufacturer’s instructions (Roche Diagnostics, Germany). More detailed information on AMH measurement has been published previously (Piltonen *et al*., 2023).

The assay limits of detection and quantitation were 0.01 ng/ml and 0.03 ng/ml for AMH. Intra-and inter-assay coefficients of variance were 1.0–1.8% and 2.9–4.4% for AMH. Limits above the measuring ranges were 23 ng/ml for AMH.

AMH concentrations were converted to pmol/l using 1 ng/ml = 7.14 pmol/l. As AMH levels are not normally distributed, for the following analysis AMH measurements were transformed using rank-based inverse normal transformation, as done previously (Ruth *et al*., 2019). For rank-based inverse normal transformation we used the package *RNOmni* in R v3.6.3.

### Genotyping details and association analysis

Genotyping of the NFBC1966 samples was performed with the Illumina Infinium HumanCNV370-Duo array. Samples were excluded based on call rate < 95%, gender mismatch, relatedness (identity by descent (IBD) estimate pihat>0.2) and outlying heterozygosity. SNPs were excluded based on missingness rate >5%, HWE p <0.000001 and MAF <1%. Genotypes were imputed with Beagle 4.1 using the SiSu v3 imputation reference panel, which consisted of 3775 individuals of Finnish ancestry with sequenced whole genomes.

Association analysis was performed using SNPTEST v2.5.4-beta1, adjusted by the first 10 genetic principal components. Markers with INFO score >0.4 were kept for the meta-analysis. Positions were converted to build hg37 to harmonise the data with the summary statistics available from Verdiesen *et al*. before running the meta-analysis, using LiftOver (Kent *et al*., 2002).

### GWAS meta-analysis

We conducted an inverse variance weighted fixed-effects meta-analysis with single genomic control correction using GWAMA software (v2.2.2) (Mägi and Morris, 2010). A total of 13,903,812 variants were included in the meta-analysis of 9,688 AMH measurements. After meta-analysis we kept variants present in both studies (totalling 7,900,839 variants) for downstream analysis. Lead SNPs were identified as SNVs independent from each other with *P*-value less than or equal to 5L×L10^−8^. The maximum distance between LD blocks of independent SNPs to merge into a single genomic locus was set to 500Lkb.

### Heritability analysis

The heritability (h^2^) was estimated by single-trait LD score regression using the meta-analysis summary statistics and HapMap 3 LD-scores using LDSC v1.0.1 (https://github.com/bulik/ldsc) (Bulik-Sullivan *et al*., 2015).

### Annotation of GWAS signals

We used FUMA (v1.4.0) to annotate the GWAS signals (Watanabe *et al*., 2017). FUMA is an online platform that performs annotation of GWAS signals using data from several databases. First, FUMA identifies lead SNPs (p-value <=5 × 10^−8^ and SNP pairwise LD r2<0.1, based on 1000G European reference) and independent significant SNPs and each risk locus (p-value <5 × 10^−8^ and LD r2< 0.6) (Supplementary Table 2). Then FUMA identifies potential candidate SNPs that are in LD with any of the identified independent significant SNPs and annotates them via linking with several databases (ANNOVAR, RegulomeDB, CADD scores etc.), which gives information on their location, functional impact, and potential regulatory effects.

### Colocalization analysis

We used HyPrColoc (Foley *et al*., 2021), a colocalisation method for identifying the overlap between our GWAS meta-analysis signals and cis-quantitative trait loci (cis-QTL) signals from different tissues and cell types (expression QTLs, transcript QTLs, exon QTLs and exon usage QTLs available in the eQTL Catalogue (Kerimov *et al*., 2021)). We lifted the GWAS summary statistics over to hg38 build to match the eQTL Catalogue using binary liftOver tool (https://genome.sph.umich.edu/wiki/LiftOver#Binary_liftOver_tool).

For the genome-wide significant (p<=5 × 10^-8^) GWAS loci identified we extracted the +/- 500kb of its top hit from QTL datasets and ran the colocalization analysis against eQTL Catalogue traits. For each eQTL Catalogue dataset we included all the QTL features which shared at least 80% of tested variants with the variants present in our GWAS region. We used the default settings for HyPrColoc analyses and did not specify any sample overlap argument, because HyPrColoc paper (Foley *et al*., 2021) demonstrates that assuming trait independence gives reasonable results. HyPrColoc outputs the posterior probability that genetic association signals for those traits are colocalising (we considered two or more signals to colocalize if the posterior probability for a shared causal variant (PP4) was 0.8 or higher). All results with a PP4 > 0.8 can be found in Supplementary Table 3.

### Gene prioritization

In order to nominate plausible candidate genes in each locus, we prioritized genes according to different levels of evidence: (1) genes containing lead variants that are coding variants or coding variants in high LD (r2L>L0.6) with lead variants; (2) genes whose expression was regulated by eQTL variants that showed significant (posterior probability >0.8) colocalization with our GWAS signals; (3) genes which showed a plausible biological role as defined previously in the research literature.

### Gene set and tissue set enrichment analysis

Gene set enrichment analysis and tissue set enrichment analysis were performed using MAGMA v1.08 implemented in FUMA v1.4.0. Gene sets were obtained from Msigcbv7.0 for ’Curated gene sets’ and ’GO terms’. A total of 15,485 gene terms were queried. The results of this analysis are presented in Supplementary Table 4 and Supplementary Figure 2. Tissue expression analysis was performed for 53 tissue types using MAGMA. The results of this analysis are presented in Supplementary Table 5 and Supplementary Figure 2.

### Genetic correlation analysis

The Complex Traits Genetics Virtual Lab (CTG-VL, https://vl.genoma.io/) was used to calculate genetic correlations between the AMH meta-analysis and 1335 traits. We applied a multiple testing correction (Benjamini-Hochberg FDR < 5%) to determine statistical significance using the *p.adjust* function in R 3.6.3. Results of the genetic correlation analysis are presented in Supplementary Figure 3 and Supplementary Table 6.

### Phenome-wide associations

We evaluated the association of our GWAS lead variants and variants in high LD with these by using human health traits and phenotypes available in the GWAS Catalog (e0_r2022-11- 29) as implemented in FUMA. We also queried the lead variants of this study from the FinnGen study (data freeze 9, total n=377,277), which combines samples collected from Finnish biobanks to digital health care data (Kurki *et al*., 2023). The results obtained are found in Supplementary Table 7 and 8 and Supplementary Figure 4.

## RESULTS

### Genome-wide association study for AMH levels

To detect genetic factors associated with circulating AMH levels, we performed a GWAS meta-analysis with data from two studies (NFBC1966 GWAS and summary statistics publicly available from Verdiesen *et al.,* 2022 including a total of 9,668 AMH measurements quantified from women of pre-menopausal age and European ancestry.

The meta-analysis identified a total of six associated loci, with six independent signals significantly associated with AMH levels (*P*□<=□5□×□10^−8^) (Table 1, Supplementary Figure 1). A total of 70 SNPs reached genome-wide significance (*P*□<=□5□×□10^−8^).

**Supplementary Figure 1.**
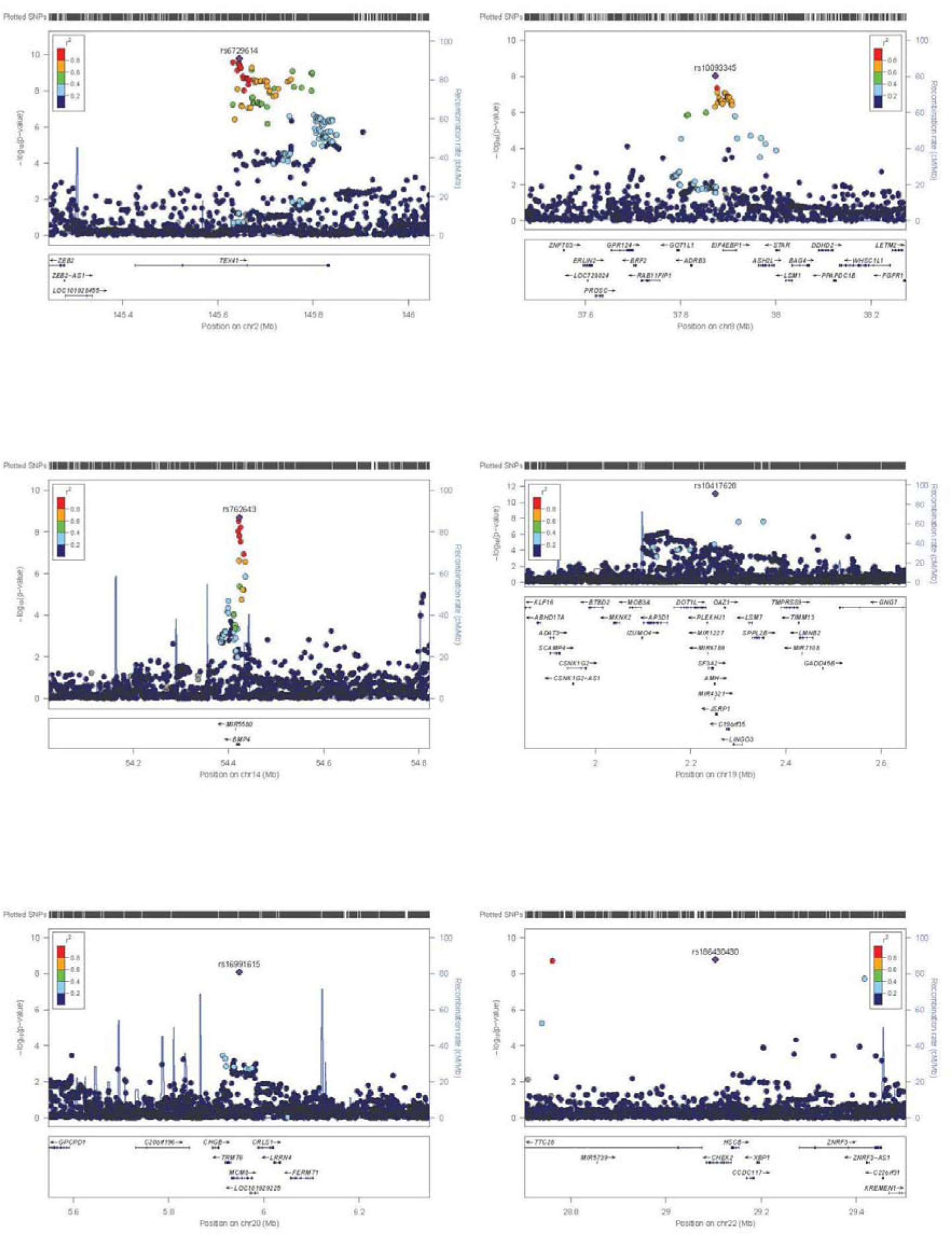
Regional plots showing the six independent loci significantly associated with AMH levels. Plots were created using the LocusZoom tool (locuszoom.org). The *y* axis represents −log_10_(*P-*values) for association of variants with AMH levels and the *x* axis represents the chromosomal positions of the variants. Each lead variant is shown as a purple diamond in each of the six genetic loci defined.

**Table 1.** **Genome-wide significant signals (p <= 5 x 10^-8^) in the meta-analysis for inverse normally transformed anti-Müllerian hormone levels in women. SNPs showing the most significant associations at each locus are shown, and novel genome-wide significant signals are highlighted in bold. EUR; European (non-Finnish), FINN: European (Finnish)**

| rsID (lead variant) | Chromosome | Position (based on build 37) | Effect allele | Effect allele frequency (EUR/FINN) | Beta (SE) | P-value | Candidate gene | Source of support for candidate gene prioritisation |
| --- | --- | --- | --- | --- | --- | --- | --- | --- |
| rs6729614 | 2 | 145644874 | G | 0.26/0.25 | 0.08 (0.01) | $5.56 \times 10^{-11}$ | *ZEB2/TEX41* | Colocalisation analysis and biological plausibility |
| **rs10093345** | **8** | **37872776** | **T** | **0.72/0.68** | **-0.08 (0.01)** | **$5.05 \times 10^{-09}$** | ***EIF4EBP1*** | **Colocalisation analysis and biological plausibility** |
| **rs762643** | **14** | **54422767** | **T** | **0.44/0.41** | **-0.07 (0.01)** | **$3.99 \times 10^{-09}$** | ***BMP4*** | **Colocalisation analysis and biological plausibility** |
| rs10417628 | 19 | 2251817 | C | 0.97/0.99 | 0.32<br>(0.04) | 9.56<br>$\times 10^{-12}$ | *AMH* | Coding variant and biological plausibility |
| rs16991615 | 20 | 5948227 | A | 0.06/0.02 | 0.16<br>(0.02) | 4.68<br>$\times 10^{-9}$ | *MCM8* | Coding variant and biological plausibility |
| **rs186430430** | **22** | **29103598** | **C** | **0.002/0.008** | **0.79**<br>**(0.12)** | **9.69**<br>**$\times 10^{-11}$** | ***CHEK2*** | **Frameshift variant c.1100delC in high LD with the lead variant** |

**Figure 1.**
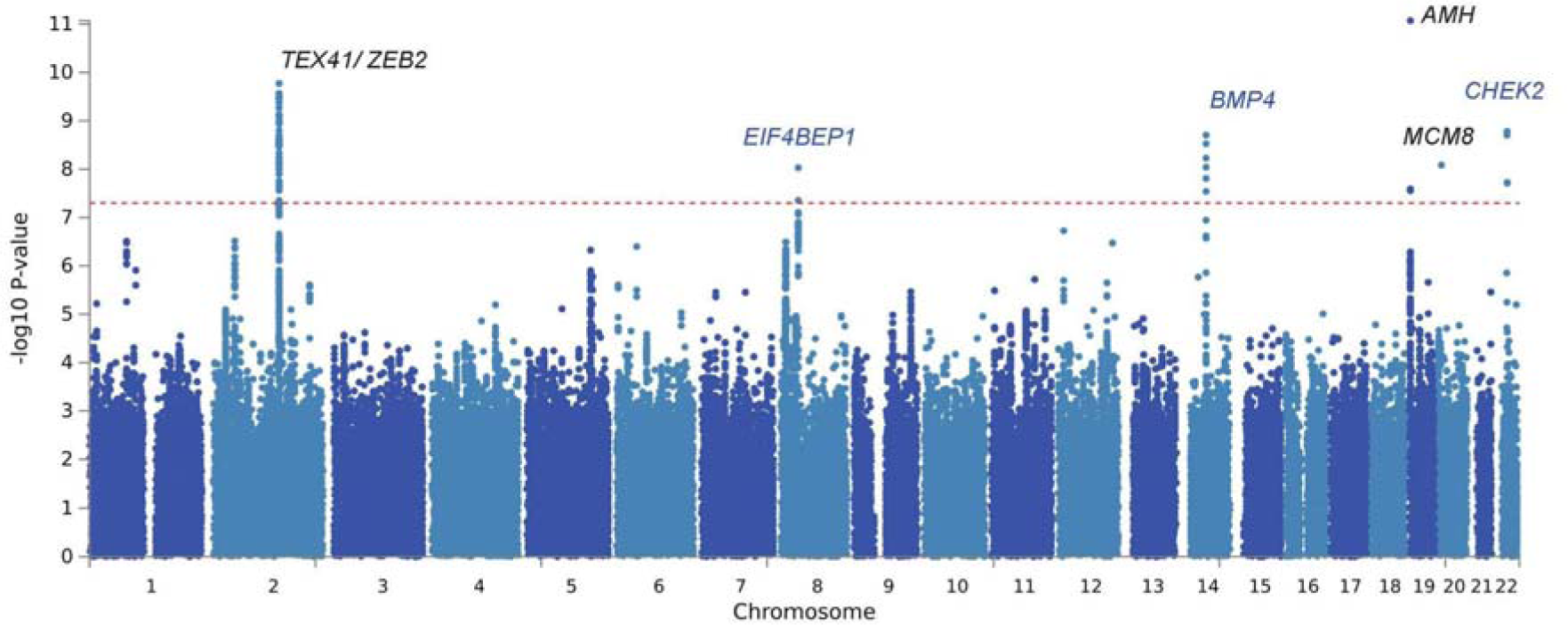
Manhattan plot for GWAS meta-analysis for AMH levels in pre-menopausal women (n=9,668). The novel loci are highlighted in blue. The *y* axis represents −log_10_(*P-* values) for association of variants with AMH levels and the *x* axis represents the chromosomal positions of the variants. The red horizontal dashed line represents the threshold for genome-wide significance (*P*□<□5□×□10^−8^).

**Figure 2.**
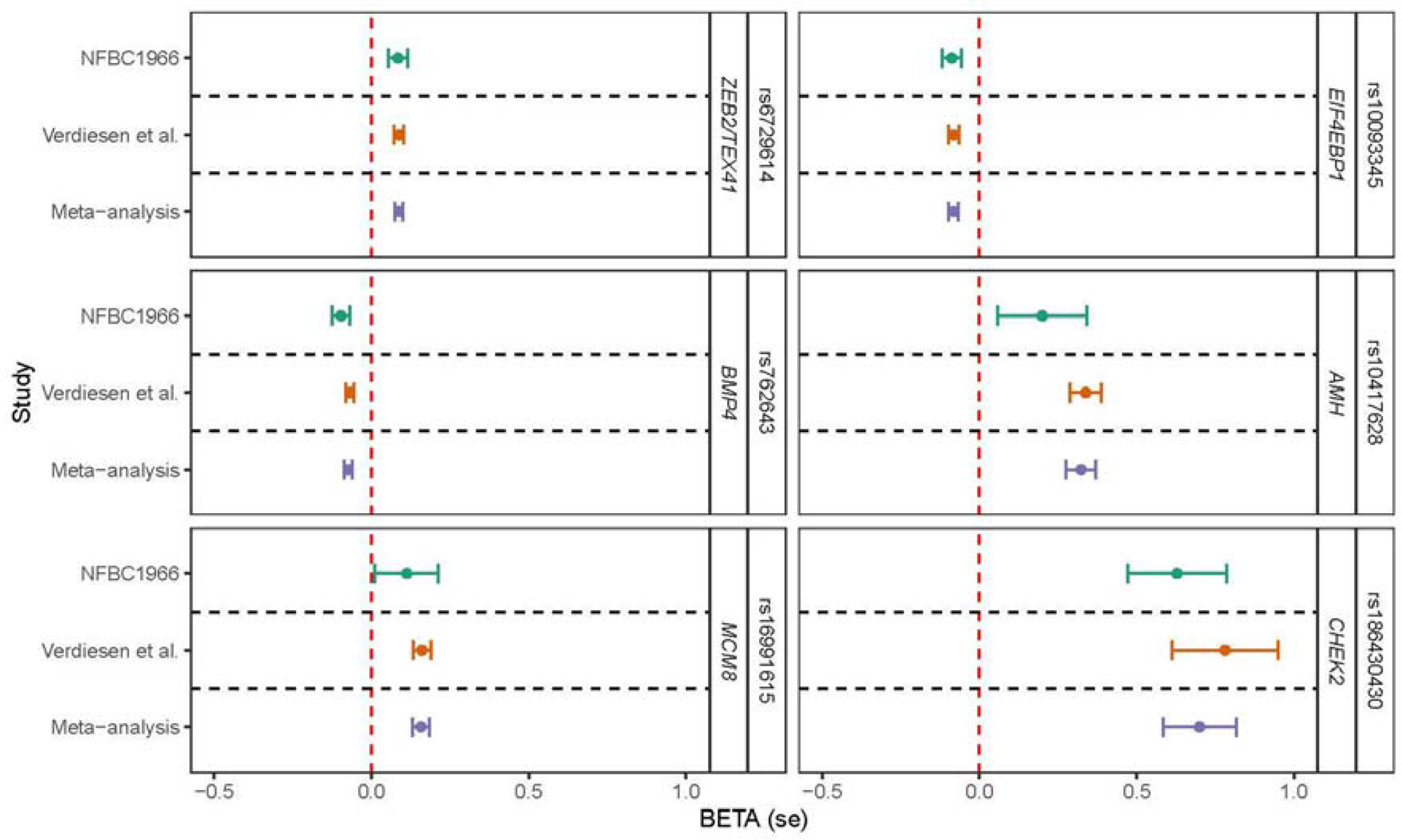
Forest plot of effect estimates for the 6 lead variants associated with AMH levels across datasets meta-analysed. The betas and standard errors are shown for the two studies meta-analysed (NFBC1966 (green dots, *n*=2,619), Verdiesen *et al*. (orange dots, *n*=7,049), and the presented largest meta-analysis (Meta-analysis, purple dots, *n*=9,668). Candidate genes are also shown next to each genetic lead signal. Betas represent the effect sizes of each genetic variant association to phenotype, assessing the same risk allele across cohorts (Effect allele and effect allele frequency are reported in Table 1).

### Characterisation of GWAS signals

Two out of the six lead variants that were detected are non-synonymous, making them notably easier to interpret and establishing a more solid foundation for pinpointing potential candidate genes. These non-synonymous variants are found in *AMH* (Anti-Mullerian Hormone) (rs10417628 in chromosome 19, p=9.56 × 10^-12^) and *MCM8* (Minichromosome Maintenance 8 Homologous Recombination Repair Factor) (rs16991615 in chromosome 20, p=4.68 × 10^-09^) and were previously described to be associated with AMH levels (Ruth *et al*., 2019; Verdiesen *et al*., 2022).

In the novel locus identified on chromosome 22, the lead variant is an intronic variant near *CHEK2* (Checkpoint Kinase 2) (rs186430430, p=9.69 × 10^-11^). Interestingly, this variant is in complete linkage disequilibrium (r^2^=1) with the frameshift variant c.1100delC in *CHEK2* (rs555607708). Despite complete linkage disequilibrium, this rare variant is only found in the association analysis of the NFBC1966 GWAS, showing a p-value of 6.69 × 10^-05^ and beta of 0.79 (s.e=0.12) in this cohort only and not found in the summary statistics results from Verdiesen *et al*., likely explained by the fact that this variant is enriched in the Finnish population (MAF= 0.008) compared to other European populations (MAF=0.002), and thus better powered to be captured in a GWAS setting including the former population in the analysis.

For the other three loci identified on chromosomes 2, 8 and 14, we defined for the first time plausible shared causal variants between GWAS signals and transcript and gene expression in specific tissues by colocalization analysis. This resulted in the prioritisation of three plausible candidate genes, respectively: *TEX41* (Testis Expressed 41), *EIF4EBP1* (Eukaryotic Translation Initiation Factor 4E Binding Protein 1) and *BMP4* (Bone Morphogenetic Protein 4) (Supplementary Table 3).

For instance, the association signal in chromosome 2, in an intronic region of *TEX41*, colocalized with eQTL signals for *TEX1* transcripts ENST00000414256 in the testis (PP4=0.99) and ENST00000445791 in the skin (PP4=1). *EIF4EBP1* was also prioritised based on colocalization evidence in three tissues, with high colocalization probability (PP4=0.90) between the GWAS signal and cis-eQTLs for *EIF4EBP1* expression in brain frontal cortex tissue, and between the GWAS signal and cis-eQTLs for *EIF4EBP1* expression in left heart ventricle. Additionally, the GWAS association signal in chromosome 8 showed colocalization with cis-eQTLs for *BMP4* transcripts ENST00000245451 and ENST00000558489 in colon transverse (PP4=0.84) and *BMP4* transcript ENST00000558489 in tibial nerve.

From colocalization results, only in the locus of chromosome 2, we observed that the associations might be explained by a single variant. Namely, we observed that the non-coding transcript (*TEX41*) exon variant rs17407477 and the intronic variant rs786244 were explaining most of the shared association, for the testis transcript expression association (posterior inclusion probability 0.90) and the skin transcript expression association (posterior inclusion probability 0.94), respectively.

Although we identified three out of the four signals that were previously reported in Verdiesen *et al*., 2022, we were unable to detect the signal near *CDCA7* described in the past work (rs11683493 (T), p=1.7 x 10^-8^). In our analysis using the NFBC66 GWAS data, this variant showed a p-value of 0.09 and therefore, the resulting p-value from the meta-analysis remained below the threshold of significance (p=1.02 x 10^-5^). Although there might be various sources of variability which explains this discrepancy in our findings, we hypothesise the difference in sample size to be the primary reason.

### Gene set and tissue set enrichment analysis

Gene set enrichment analysis highlighted significant enrichment of renal system vasculature morphogenesis (p=5.2×10^-8^), glomerulus vasculature morphogenesis (p=1.77×10^-7^) and glomerulus morphogenesis (p=2.99×10^-6^) (see Supplementary Table 4 and Supplementary Figure 2). Tissue expression analysis yielded the strongest enrichment in the pituitary gland, even though not reaching significance after multiple testing correction (p_non-adj_=0.02) (see Supplementary Figure 2 and Supplementary Table 5 and Supplementary Figure 2).

**Supplementary Figure 2.**
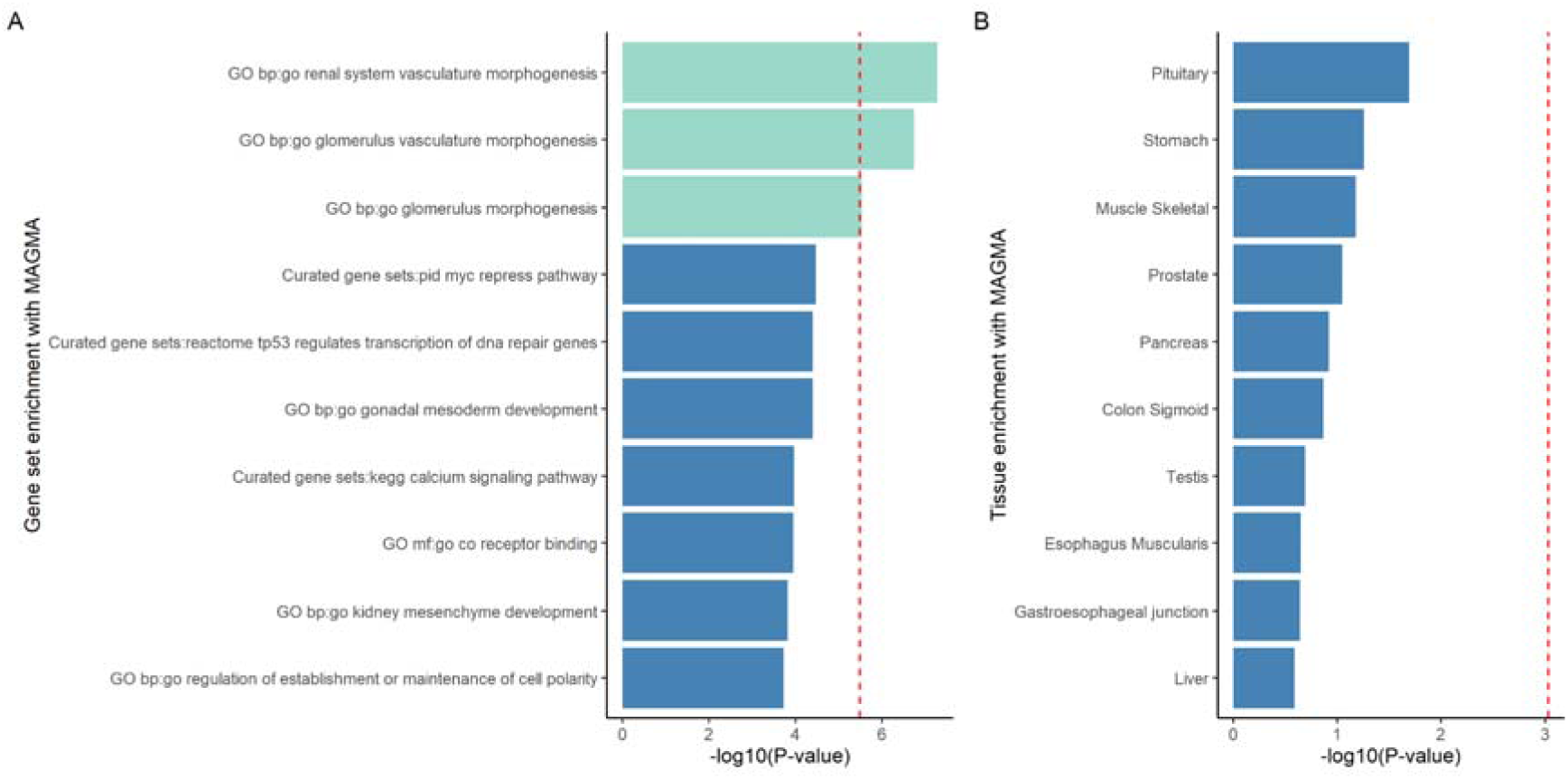
Gene set and tissue set enrichment results. **A) Gene set enrichment analyses with MAGMA.** Green bars and the red dashed line indicate Bonferroni threshold, set to p=0.05/15485=3.22 x 10^-6^. **B) Tissue enrichment analyses with MAGMA.** Red dashed line indicates Bonferroni threshold, set to p=0.05/54=0.0009.

### Genetic correlations of AMH levels with diseases and traits

From genetic correlation analysis assessing the relationship between AMH levels and 1,335 traits (summary statistics for those traits are found in http://www.nealelab.is/uk-biobank/), three traits reflecting menopausal timing remained significant after FDR correction, all displaying very high genetic correlations with AMH levels: ‘Age at menopause’ (r_g_=0.95, se=0.21, p=8.85 x 10^-6^), ‘Had menopause’ (referring to having already undergone menopause) (r_g_=-0.99, se=0.22, p=1.56 x 10^-5^) and ‘Age started hormone-replacement therapy (HRT)’ (age for initiation of hormone replacement therapy) (r_g_=0.92, se=0.21, p=2.05 x 10^-5^). Additionally, we observed a range of nominally significant associations (p<0.05) with traits spanning reproductive/hormonal traits (such as polyp of female genital tract and female genital prolapse), metabolic traits such as inverse relationship with HbA1c and positive correlations with neoplasms such as breast and bowel cancer. (Supplementary Figure 3 and full results and details can be found in Supplementary Table 6).

**Supplementary Figure 3.**
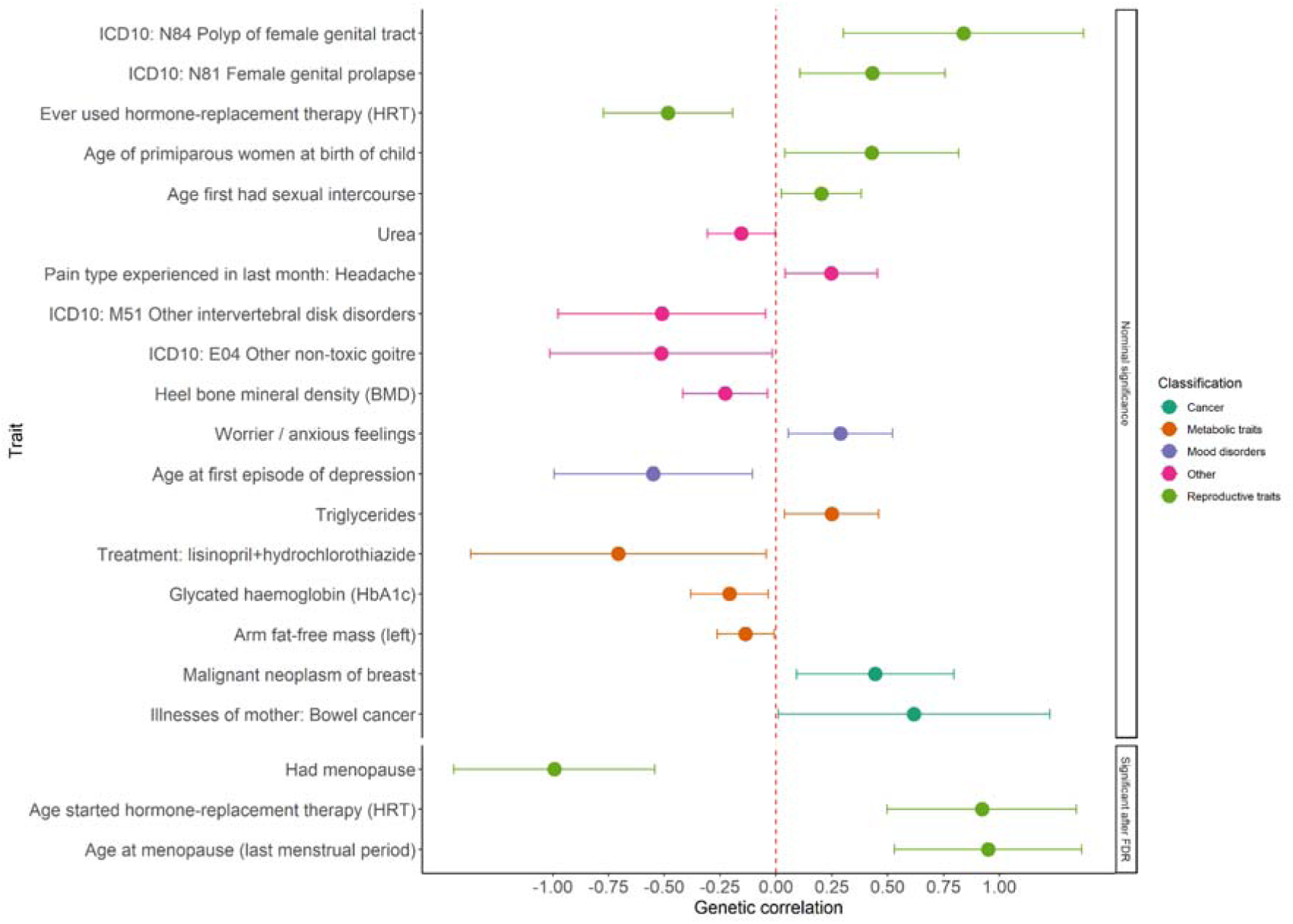
Genetic correlation results between AMH summary statistics and publicly available summary statistics for other traits. Out of 1,335 associations, the lower section depicts significant associations after FDR correction (p-adj<0.05) and the upper section depicts nominal significant correlations (p<0.05). Dots show the estimated genetic correlation (rg) and error bars indicate 95% confidence limits. Dotted red line indicates no genetic correlation. ICD10: International Classifications of Diseases 10th Revision.

### Phenome-wide association study

To get more insights into phenotypic manifestations of the AMH-associated variants, we performed a phenome-wide association study using the GWAS Catalog. We detected that associated variants in the six loci identified had associations across several traits, spanning from cardiovascular traits, age at menopause, estradiol levels, heel bone mineral density, breast cancer, uterine fibroids, PCOS to blood cell counts (Supplementary Table 7 and Supplementary Figure 3).

We performed a further look-up of the lead variants in the FinnGen study (data freeze 9, total n=377,277), which combines samples collected from Finnish biobanks to digital health care data. We detected that two of the lead variants had associations (p<1 x 10^-5^) with outcomes in FinnGen, rs762643 (near *BMP4*) associated with cervical disc disorders and hernia, while rs186430430 (near *CHEK2*) associated with a variety of outcomes including diseases of the genitourinary tract, different neoplasms (e.g. leiomyoma of uterus, neoplasms of ovary), and breast-cancer-related outcomes (full results can be found in Supplementary Table 8).

**Supplementary Figure 4.**
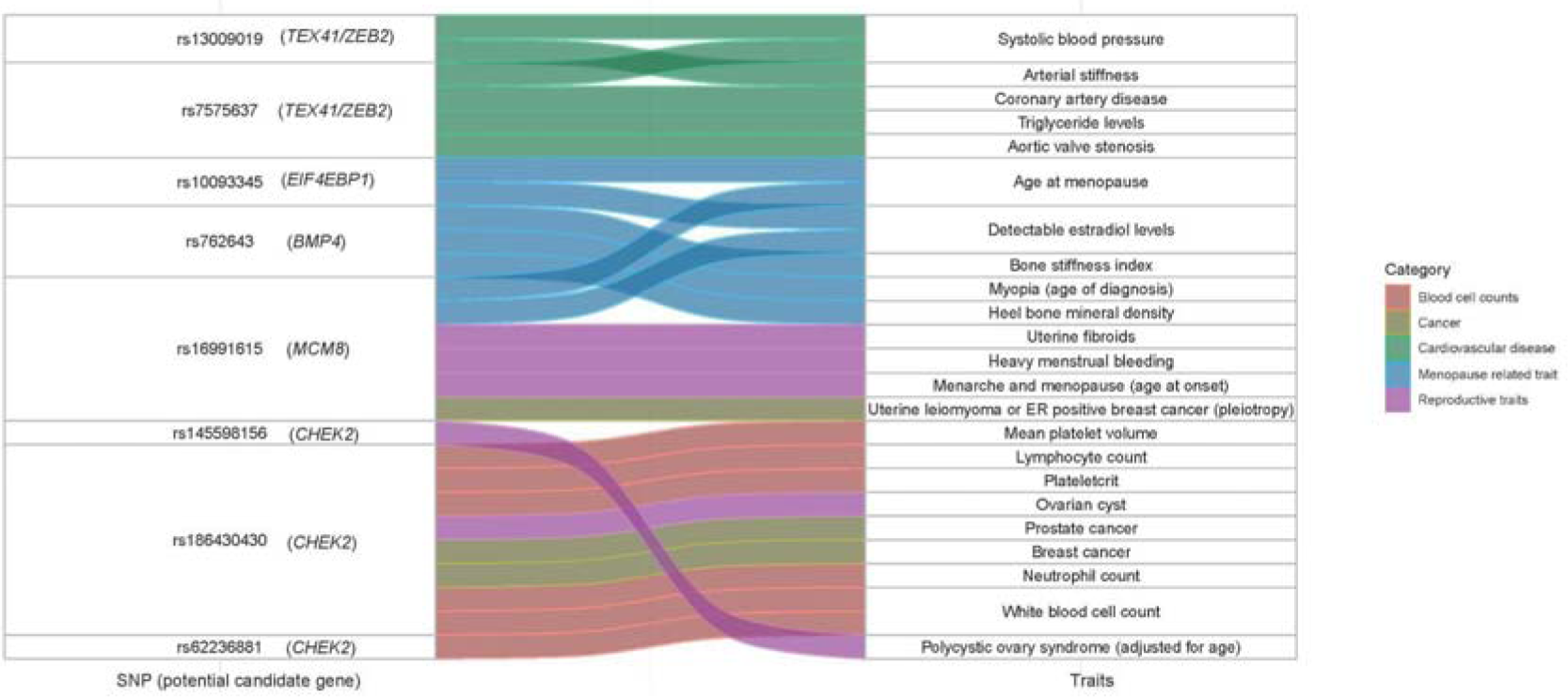
**River plot showing the association between lead variants (or variants in high LD with these) identified in our AMH GWAS (left column) and independent traits which show significant association results for respective variants in the GWAS Catalog (right column).**

## DISCUSSION

Our GWAS meta-analysis of circulating AMH measurements in 9,668 pre-menopausal women, including 2,619 measurements from women at age 31 years old from the NFBC1966, identified three novel association signals near *EIF4EBP1*, *BMP4* and *CHEK2*. We also detected three previously identified signals near *TEX41*, *MCM8*, and *AMH* (Verdiesen *et al*., 2022).

*CHEK2* was one of the most interesting associated loci. Of note, the lead variant near *CHEK2* (rs186430430, p=9.69 × 10^-11^, MAF= 0.002) is in complete linkage disequilibrium (r^2^=1) with a frameshift variant, c.1100delC (rs555607708, found in NFBC GWAS only, p=6.69 × 10^-05^, OR=1.87, 95% CI= 1.37-2.55), as it is enriched in the Finnish population (MAF=0.008) compared to other non-Finnish and non-Estonian European populations (MAF=0.002) (Tyrmi *et al*., 2022). Apart from being known as a moderate risk gene for breast cancer, different GWA studies highlighted loss of function alleles in *CHEK2* to be associated with firstly, PCOS (Tyrmi *et al*., 2021) and secondly, with ovarian ageing (assessed as age at natural menopause) (Ruth *et al*., 2021). Furthermore, recent research done with exome data has also identified an association between *CHEK2* truncating variants (excluding c.1110delC) and later age at menarche (Kentistou *et al*., 2023).

*CHEK2* encodes a checkpoint kinase 2, which induces cell cycle arrest and apoptosis in response to DNA damage (Ahn *et al*., 2004), also in oocytes with unrepaired DNA damage (Ruth *et al*., 2021). In the work from *Ruth et al.,* 2021 Chek2-/- female mice showed reduced follicular atresia around reproductive senescence, increased follicular response to gonadotrophin stimulation, and also elevated AMH levels around reproductive senescence. Our finding goes in line with this research and supports the hypothesis that loss of function in *CHEK2* might result in decreased follicular atresia and higher AMH levels also in young premenopausal women.

In our general meta-analysis, we also detected previously identified associations of a missense variant in *MCM8* (Ruth *et al*., 2019; Verdiesen *et al*., 2022), a known DNA repair gene (Park *et al*., 2013). This missense variant has been associated previously with premature ovarian failure, infertility, age at menopause (Day *et al*., 2015) and cancer (Michailidou *et al*., 2017; Griffin and Trakselis, 2019; Lutzmann *et al*., 2019). However, this association was the weakest in NFBC66 alone. While this could support an age-specific effect of the missense variant, as supported in Verdiesen *et al*. upon identifying a negative effect direction for the cohort including adolescents versus older cohorts, this discrepancy might be as well due to the variant’s lower frequency in the Finnish population (MAF=0.02 compared to MAF=0.06 in other European populations) and a smaller sample size in the NFBC1966 study. Further research including age-stratified groups with AMH levels available and genetic data might inform potential age-specific effects of genetic signals.

Regarding the missense variant in the AMH gene also reported in Verdiesen *et al*., the association between the missense variant rs10417628 in the *AMH* gene and AMH levels remains unclear in the current analysis. A case report from 2020 suggested that this variant reduces AMH detection without affecting its bioactivity (Hoyos *et al*., 2020). Previous GWAS meta-analysis also provided inconclusive results due to inconsistent evidence from different assay data. In the NFBC66 study using an automated assay (Elecsys® AMH Plus), the missense variant showed a p-value of 0.15 (n=2,619), supporting the hypothesis of a detection issue rather than an actual difference in AMH bioactivity. However, comparing AMH values from different assays remains problematic, and recent evidence suggests lower values in automated assays compared to Gen II and Ansh Labs assay (Moolhuijsen and Visser, 2020). Further research is needed to validate the association between rs10417628 and AMH levels using different assays.

Colocalization analysis supports regulatory effects of GWAS variants associated with AMH with genetic variants modifying expression levels of *TEX41*, *BMP4* and *EIF4EBP1* and thus provides a refinement for the characterization of possible regulatory effects of the genetic variants on different transcripts and genes.

Interestingly, we observed high posterior probability (PP4=0.99) of colocalization between our GWAS signal and eQTL signal for the *TEX41* (Testis expressed 41) transcript ENST00000414256 in the testis, prioritising this long non-coding RNA gene for the first time in association with AMH levels based on colocalization analysis. In the same locus*, ZEB2* shows biological plausibility as a candidate gene since it plays a role as inhibitor of signal transduction in TGF-B and BMP signalling through interaction with ligand-activated SMAD proteins (Postigo *et al*., 2003; Conidi *et al*., 2011).

Colocalization analysis also supported the nomination of *EIF4EBP1* in the novel locus identified in chromosome 8. This gene encodes a translation repressor protein that competitively binds to eukaryotic translation initiation factor 4E (EIF4E) (Gingras *et al*., 1999; Harris and Lawrence, 2003) and is a major substrate of mTOR and a key player in mTOR signalling pathway. Studies show that both EIF4EBP1 and EIF4E are involved in cancer development and progression where up-regulated EIF4E plays an oncogenic role in carcinogenesis (Heikkinen *et al*., 2013; Cha *et al*., 2015). The same intergenic lead variant in this locus has been associated with age at menopause, and other menopause-related traits (ever had menopause and ever used hormone-replacement therapy), supporting a plausible association with AMH levels.

We observed colocalization between our GWAS signal and variants modulating gene expression of *BMP4*. *BMP4* has a known regulatory role on AMH expression through activation of the SMAD proteins (Estienne *et al*., 2015; Pierre *et al*., 2016). Smad interacting protein 1 (also known as *ZEB2)* is one of the plausible candidate genes in chromosome 2, which would as well interact with SMAD proteins and participate in AMH regulation (Postigo *et al*., 2003).

Our GWAS of AMH measurements revealed significant enrichment in renal system vascular morphogenesis, glomerulus vascular morphogenesis, and glomerulus morphogenesis gene set analysis. This supports the close connection between urinary and reproductive system development, originating from a common embryological origin, the intermediate mesoderm. Additionally, in our gene set enrichment results, we observed a nominal significance association with the gonadal mesoderm gene set (p=4.2 × 10^-5^), further supporting the interconnectedness of the urinary and reproductive systems during development. We propose that this observation may be linked to the identification of *BMP4* locus associated with AMH levels. The BMP4 signalling pathway is crucial during kidney development, including ureteric bud outgrowth (Grinspon and Rey, 2014; Nishinakamura and Sakaguchi, 2014; Oxburgh *et al*., 2014). This observation suggests that genetic variants affecting the renal system’s development and function could influence AMH levels in women later in life. To gain further insight, studying AMH levels in younger cohorts and conducting sex-stratified analyses may be valuable in understanding the association between urogenital development and AMH levels in postnatal stages.

Tissue expression analysis also identified enrichment in the pituitary gland, although this did not reach significance after multiple testing correction. This result goes in line with research from recent years showing that AMH has versatile actions in different levels of the hypothalamus-pituitary-gonadal axis (Silva and Giacobini, 2021), further supporting the developmental alterations of neuroendocrine circuits regulating fertility.

We identified three significant genetic correlations which align with the observations in Verdiesen *et al*., indicating a strong positive genetic correlation between AMH and traits reflecting age at menopause, suggesting shared underlying genetic risk factors and supporting AMH as a proxy for ovarian reserve. We also found interesting nominal significant associations, pointing towards a shared genetic background between AMH levels and breast cancer, consistent with epidemiological studies (Ge *et al*., 2018). We hypothesise both *MCM8* and *CHEK2* loci may play a role in this relationship and Mendelian randomisation studies with independent samples could further investigate this association’s direction.

Upon querying the AMH GWAS lead variants and variants in high linkage disequilibrium with those in other traits, we found shared significant signals amongst known epidemiological associations (Homburg and Crawford, 2014; De Kat *et al*., 2017; Ge *et al*., 2018; Moolhuijsen and Visser, 2020) with AMH such as PCOS, breast cancer, postmenopausal status (age at menopause, estradiol levels, heel bone mineral density) and cardiovascular disease. Additionally, we observed novel associations with other traits in European ancestry studies, including uterine fibroids (coming from signal in *MCM8*) and blood cell counts (coming from signals near *CHEK2*, which has been also associated recently with clonal hematopoiesis (Kar *et al*., 2022). Further epidemiological studies assessing the potential causal relationships between these traits and AMH levels are warranted to better understand the mechanisms underlying these associations.

Our study has several strengths, including a large sample size and inclusion of a founder population, which allowed us to identify novel rare variants associated with AMH levels. Furthermore, a younger cohort and equal age of measurement from 2,619 women increases the variability of AMH levels and boosts the power to detect associations. We also performed colocalization analysis for the first time, which provides a refinement for the characterization of possible regulatory effects of the genetic variants on different transcripts and genes and detect significant gene set enrichment. Our study has some limitations to consider. First, we only included women of European ancestry, and our findings may not be generalizable to other populations. Second, as with other reproductive phenotypes, the lack of sufficiently sized datasets from relevant tissue in commonly used gene expression databases hinders a more reliable assessment of the mechanisms underlying regulatory effects in reproductive tissues.

In conclusion, our study expands our understanding of the genetic determinants of serum AMH levels in pre-menopausal women by identifying new loci associated with serum AMH concentration. Our results highlight the increased power of founder populations and larger sample sizes to boost the discovery of novel trait-associated variants underlying variation in AMH levels and to explore plausible genetic regulatory effects of the variants identified. Further studies are needed to validate our findings and to explore the biological mechanisms underlying the identified associations.

## Data Availability

The GWAS meta-analysis summary statistics will be available for download from the GWAS Catalog. Accession numbers will be provided upon publication.

## Acknowledgments

We want to thank the participants and investigators of the FinnGen study and the NFBC1966 study. Part of the computations were performed in the High-Performance Computing Center of University of Tartu.

## Bibliography

Ahn J, Urist M, Prives C. The Chk2 protein kinase. DNA Repair (Amst) [Internet] 2004;3:1039–1047. DNA Repair (Amst).

Bulik-Sullivan B, Loh PR, Finucane HK, Ripke S, Yang J, Patterson N, Daly MJ, Price AL, Neale BM, Corvin A, et al. LD score regression distinguishes confounding from polygenicity in genome-wide association studies. Nat Genet 2015;47:291–295. Nature Publishing Group.

Cha YL, Li PD, Yuan LJ, Zhang MY, Zhang YJ, Rao HL, Zhang HZ, Zheng XFS, Wang HY. EIF4EBP1 overexpression is associated with poor survival and disease progression in patients with hepatocellular carcinoma. PLoS One [Internet] 2015;10:. PLoS One.

Conidi A, Cazzola S, Beets K, Coddens K, Collart C, Cornelis F, Cox L, Joke D, Dobreva MP, Dries R, et al. Few Smad proteins and many Smad-interacting proteins yield multiple functions and action modes in TGFβ/BMP signaling in vivo. Cytokine Growth Factor Rev [Internet] 2011;22:287–300. Cytokine Growth Factor Rev.

Day FR, Ruth KS, Thompson DJ, Lunetta KL, Pervjakova N, Chasman DI, Stolk L, Finucane HK, Sulem P, Bulik-Sullivan B, et al. Large-scale genomic analyses link reproductive aging to hypothalamic signaling, breast cancer susceptibility and BRCA1-mediated DNA repair. Nat Genet [Internet] 2015;47:1294–1303. Nat Genet.

Estienne A, Pierre A, Clemente N Di, Picard JY, Jarrier P, Mansanet C, Monniaux D, Fabre S. Anti-Müllerian hormone regulation by the bone morphogenetic proteins in the sheep ovary: deciphering a direct regulatory pathway. Endocrinology [Internet] 2015;156:301–313. Endocrinology.

Finkelstei JS, Lee H, Karlamangla A, Nee RM, Slus PM, Burnett-Bowie SAM, Darakananda K, Donaho PK, Harlo SD, Prizan SH, et al. Antimullerian Hormone and Impending Menopause in Late Reproductive Age: The Study of Women’s Health Across the Nation. J Clin Endocrinol Metab [Internet] 2020;105:E1862–E1871. J Clin Endocrinol Metab.

Foley CN, Staley JR, Breen PG, Sun BB, Kirk PDW, Burgess S, Howson JMM. A fast and efficient colocalization algorithm for identifying shared genetic risk factors across multiple traits. Nat Commun [Internet] 2021;12:. Nat Commun.

Ge W, Clendenen T V., Afanasyeva Y, Koenig KL, Agnoli C, Brinton LA, Dorgan JF, Eliassen AH, Falk RT, Hallmans G, et al. Circulating anti-Müllerian hormone and breast cancer risk: A study in ten prospective cohorts. Int J cancer [Internet] 2018;142:2215– 2226. Int J Cancer.

Gingras AC, Raught B, Sonenberg N. eIF4 initiation factors: effectors of mRNA recruitment to ribosomes and regulators of translation. Annu Rev Biochem 1999;68:913–963. United States.

Griffin WC, Trakselis MA. The MCM8/9 complex: A recent recruit to the roster of helicases involved in genome maintenance. DNA Repair (Amst) [Internet] 2019;76:1–10. DNA Repair (Amst).

Grinspon RP, Rey RA. When hormone defects cannot explain it: malformative disorders of sex development. Birth Defects Res C Embryo Today [Internet] 2014;102:359–373. Birth Defects Res C Embryo Today.

Harris TE, Lawrence JCJ. TOR signaling. Sci STKE 2003;2003:re15. United States.

Heikkinen T, Korpela T, Fagerholm R, Khan S, Aittomäki K, Heikkilä P, Blomqvist C, Carpén O, Nevanlinna H. Eukaryotic translation initiation factor 4E (eIF4E) expression is associated with breast cancer tumor phenotype and predicts survival after anthracycline chemotherapy treatment. Breast Cancer Res Treat [Internet] 2013;141:79–88. Breast Cancer Res Treat.

Homburg R, Crawford G. The role of AMH in anovulation associated with PCOS: a hypothesis. Hum Reprod [Internet] 2014;29:1117–1121. Hum Reprod.

Hoyos LR, Visser JA, McLuskey A, Chazenbalk GD, Grogan TR, Dumesic DA. Loss of anti-Müllerian hormone (AMH) immunoactivity due to a homozygous AMH gene variant rs10417628 in a woman with classical polycystic ovary syndrome (PCOS). Hum Reprod [Internet] 2020;35:2294–2302. Hum Reprod.

Kar SP, Quiros PM, Gu M, Jiang T, Mitchell J, Langdon R, Iyer V, Barcena C, Vijayabaskar MS, Fabre MA, et al. Genome-wide analyses of 200,453 individuals yield new insights into the causes and consequences of clonal hematopoiesis. Nat Genet [Internet] 2022;54:1155–1166. Nat Genet.

Kat AC De, Monique Verschuren W, Eijkemans MJC, Broekmans FJM, Schouw YT Van Der. Anti-Müllerian Hormone Trajectories Are Associated With Cardiovascular Disease in Women: Results From the Doetinchem Cohort Study. Circulation [Internet] 2017;135:556–565. Circulation.

Kent WJ, Sugnet CW, Furey TS, Roskin KM, Pringle TH, Zahler AM, Haussler and D. The human genome browser at UCSC. Genome Res [Internet] 2002;12:996–1006. Genome Res.

Kentistou KA, Kaisinger LR, Stankovic S, Vaudel M, Oliveira EM de, Messina A, Walters RG, Liu X, Busch AS, Helgason H, et al. Understanding the genetic complexity of puberty timing across the allele frequency spectrum. medRxiv [Internet] 2023; Cold Spring Harbor Laboratory Press Available from: https://www.medrxiv.org/content/early/2023/06/20/2023.06.14.23291322.

Kerimov N, Hayhurst JD, Peikova K, Manning JR, Walter P, Kolberg L, Samoviča M, Sakthivel MP, Kuzmin I, Trevanion SJ, et al. A compendium of uniformly processed human gene expression and splicing quantitative trait loci. Nat Genet [Internet] 2021;53:1290–1299.

Kurki MI, Karjalainen J, Palta P, Sipilä TP, Kristiansson K, Donner KM, Reeve MP, Laivuori H, Aavikko M, Kaunisto MA, et al. FinnGen provides genetic insights from a well-phenotyped isolated population. Nature [Internet] 2023;613:508–518. Nature.

Lutzmann M, Bernex F, Costa de Jesus C da, Hodroj D, Marty C, Plo I, Vainchenker W, Tosolini M, Forichon L, Bret C, et al. MCM8- and MCM9 Deficiencies Cause Lifelong Increased Hematopoietic DNA Damage Driving p53-Dependent Myeloid Tumors. Cell Rep [Internet] 2019;28:2851–2865.e4. Cell Rep.

Mägi R, Morris AP. GWAMA: software for genome-wide association meta-analysis. BMC Bioinformatics 2010;11:288.

Michailidou K, Lindström S, Dennis J, Beesley J, Hui S, Kar S, Lemaçon A, Soucy P, Glubb D, Rostamianfar A, et al. Association analysis identifies 65 new breast cancer risk loci. Nature [Internet] 2017;551:92–94. Nature.

Moolhuijsen LME, Visser JA. Anti-Müllerian Hormone and Ovarian Reserve: Update on Assessing Ovarian Function. J Clin Endocrinol Metab [Internet] 2020;105:. J Clin Endocrinol Metab.

Nishinakamura R, Sakaguchi M. BMP signaling and its modifiers in kidney development. Pediatr Nephrol [Internet] 2014;29:681–686. Pediatr Nephrol.

Nordström T, Miettunen J, Auvinen J, Ala-Mursula L, Keinänen-Kiukaanniemi S, Veijola J, Järvelin M-R, Sebert S, Männikkö M. Cohort Profile: 46 years of follow-up of the Northern Finland Birth Cohort 1966 (NFBC1966). Int J Epidemiol 2022;50:1786–1787j. England.

Oxburgh L, Brown AC, Muthukrishnan SD, Fetting JL. Bone morphogenetic protein signaling in nephron progenitor cells. Pediatr Nephrol [Internet] 2014;29:531–536. Pediatr Nephrol.

Park J, Long DT, Lee KY, Abbas T, Shibata E, Negishi M, Luo Y, Schimenti JC, Gambus A, Walter JC, et al. The MCM8-MCM9 complex promotes RAD51 recruitment at DNA damage sites to facilitate homologous recombination. Mol Cell Biol [Internet] 2013;33:1632–1644. Mol Cell Biol.

Pierre A, Estienne A, Racine C, Picard JY, Fanchin R, Lahoz B, Alabart JL, Folch J, Jarrier P, Fabre S, et al. The Bone Morphogenetic Protein 15 Up-Regulates the Anti-Müllerian Hormone Receptor Expression in Granulosa Cells. J Clin Endocrinol Metab [Internet] 2016;101:2602–2611. J Clin Endocrinol Metab.

Piltonen TT, Komsi E, Morin-Papunen LC, Korhonen E, Franks S, Järvelin M-R, Arffman RK, Ollila M-M. AMH as part of the diagnostic PCOS workup in large epidemiological studies. Eur J Endocrinol 2023;188:547–554. England.

Postigo AA, Depp JL, Taylor JJ, Kroll KL. Regulation of Smad signaling through a differential recruitment of coactivators and corepressors by ZEB proteins. EMBO J [Internet] 2003;22:2453–2462. EMBO J.

Ruth KS, Day FR, Hussain J, Martínez-Marchal A, Aiken CE, Azad A, Thompson DJ, Knoblochova L, Abe H, Tarry-Adkins JL, et al. Genetic insights into biological mechanisms governing human ovarian ageing. Nature [Internet] 2021;596:393–397. Nature.

Ruth KS, Soares ALG, Borges MC, Eliassen AH, Hankinson SE, Jones ME, Kraft P, Nichols HB, Sandler DP, Schoemaker MJ, et al. Genome-wide association study of anti-Müllerian hormone levels in pre-menopausal women of late reproductive age and relationship with genetic determinants of reproductive lifespan. Hum Mol Genet [Internet] 2019;28:1392–1401. Hum Mol Genet.

Schuh-Huerta SM, Johnson NA, Rosen MP, Sternfeld B, Cedars MI, Reijo Pera RA. Genetic markers of ovarian follicle number and menopause in women of multiple ethnicities. Hum Genet [Internet] 2012;131:1709–1724. Hum Genet.

Silva MSB, Giacobini P. New insights into anti-Müllerian hormone role in the hypothalamic-pituitary-gonadal axis and neuroendocrine development. Cell Mol Life Sci [Internet] 2021;78:. Cell Mol Life Sci.

Tyrmi JS, Arffman RK, Pujol-Gualdo N, Kurra V, Morin-Papunen L, Sliz E, FinnGen, Team EBR, Piltonen TT, Laisk T, et al. Leveraging Northern European population history; novel low frequency variants for polycystic ovary syndrome. medRxiv [Internet] 2021;2021.05.20.21257510Available from: http://medrxiv.org/content/early/2021/05/24/2021.05.20.21257510.abstract.

Tyrmi JS, Arffman RK, Pujol-Gualdo N, Kurra V, Morin-Papunen L, Sliz E, Piltonen TT, Laisk T, Kettunen J, Laivuori H. Leveraging Northern European population history: novel low-frequency variants for polycystic ovary syndrome. Hum Reprod [Internet] 2022;37:352–365. Hum Reprod.

Verdiesen RMG, Schouw YT Van Der, Gils CH Van, Verschuren WMM, Broekmans FJM, Borges MC, Gonçalves Soares AL, Lawlor DA, Eliassen AH, Kraft P, et al. Genome-wide association study meta-analysis identifies three novel loci for circulating anti-Müllerian hormone levels in women. Hum Reprod [Internet] 2022;37:1069–1082. Oxford University Press.

Watanabe K, Taskesen E, Bochoven A van, Posthuma D. Functional mapping and annotation of genetic associations with FUMA. Nat Commun 2017;8:1826.

Weenen C, Laven JSE, Bergh ARM von, Cranfield M, Groome NP, Visser JA, Kramer P, Fauser BCJM, Themmen APN. Anti-Müllerian hormone expression pattern in the human ovary: potential implications for initial and cyclic follicle recruitment. Mol Hum Reprod [Internet] 2004;10:77–83. Mol Hum Reprod.

